# Asthma inflammatory phenotypes on four continents: most asthma cases are non-eosinophilic

**DOI:** 10.1101/2020.08.23.20177162

**Authors:** Lucy Pembrey, Collin Brooks, Harriet Mpairwe, Camila A Figueiredo, Aida Y. Oviedo, Martha Chico, Hajar Ali, Irene Nambuya, Pius Tumwesige, Steven Robertson, Susan Ring, Mauricio L. Barreto, Philip J. Cooper, John Henderson, Alvaro A. Cruz, Jeroen Douwes, Neil Pearce, the WASP Study Group

**Affiliations:** London School of Hygiene and Tropical Medicine; Centre for Public Health Research, Massey University, Wellington, New Zealand; MRC/UVRI and LSHTM Uganda Research Unit, Entebbe, Uganda; Institute of Collective Health, Federal University of Bahia, Salvador, Brazil; Fundacion Ecuatoriana Para Investigacion en Salud, Quito, Ecuador; Population Health Sciences, Bristol Medical School, University of Bristol, United Kingdom; MRC Integrative Epidemiology Unit at University of Bristol, Bristol BS8 2BN, United Kingdom; Center for Data and Knowledge Integration for Health (CIDACS), Fiocruz, Bahia; Universidad Internacional del Ecuador, Quito, Ecuador; St George’s University of London, United Kingdom; ProAR, Federal University of Bahia, Salvador, Brazil; Institute for Health Sciences, Federal University of Bahia, Salvador, Brazil

## Abstract

**Rationale:** In high-income countries less than half of asthma cases involve eosinophilic airways inflammation. Few studies have measured inflammatory asthma phenotypes in low-and-middle-income countries.

**Objectives:** To assess asthma inflammatory phenotypes in Brazil, Ecuador, Uganda, New Zealand and the United Kingdom.

**Methods:** We recruited 998 asthmatics and 356 non-asthmatics: 204/40 in Brazil, 176/67 in Ecuador, 235/132 in New Zealand, 207/50 in Uganda, and 176/67 in the United Kingdom. All centres studied children and adolescents (age-range 8-20 years), except the UK centre involved 26-27 year olds. Information was collected using questionnaires, clinical characterisation, blood, and induced sputum.

**Measurements and main results:** 87% provided a sputum sample, ranging from 74% (Brazil) to 93% (United Kingdom); of these, 71% were countable. The proportion of asthmatics classified as eosinophilic asthma (EA) was 39% (95% confidence interval 35%-43%) overall: 35% in Brazil, 32% in Ecuador, 50% in New Zealand, 33% in Uganda, and 33% in the United Kingdom. The non-eosinophilic asthmatics (NEA) had similar severity/control to the eosinophilic asthmatics (EA). Of the 61% (95%CI 57%-65%) of cases with NEA, 50% showed no signs of inflammation (paucigranulocytic), with 11% having neutrophilic inflammation.

**Conclusions:** This is the first time that sputum induction has been used to compare asthma inflammatory phenotypes in high-income countries and low-and-middle-income countries. Most cases were non-eosinophilic. This has major implications for asthma prevention and management in both of these contexts, and supports the need to recognise asthma as a heterogeneous condition, and to develop new prevention strategies and new therapies which target NEA.

## Introduction

It is now well established in high-income countries (HICs) that there are multiple phenotypes and endotypes of asthma ^1, 2^, and that generally less than one half of asthma cases are attributable to eosinophilic airways inflammation^3, 4^. Most asthma in low- and middle-income countries (LMICs) appears to be non-atopic^5^, the association between atopy and asthma is much weaker than in high income countries (HICs)^5, 6^, and standardised comparisons across populations or time periods also show only weak and inconsistent associations between the prevalence of atopy and the prevalence of asthma^5^. However, these comparisons have generally involved skin-prick-test positivity or allergen-specific IgE^7^, and few studies in LMICs have measured inflammatory asthma phenotypes directly.

As early as 1958 it was known that non-eosinophilic asthma (NEA) occurred and was not responsive to systemic corticosteroids, but this evidence was largely ignored until the last 20 years.^8^ A recent Lancet Commission report on asthma called for greater recognition of the various phenotypes with different underlying pathological mechanisms, often grouped under the non-specific label of asthma^9^. The importance of non-eosinophilic asthma in particular was highlighted at a recent Academia Europea/Asthma UK meeting^10^, with the meeting report noting non-eosinophilic asthma “responds only poorly to the long-standing, conventional treatments traditionally used to reduce inflammation of the airways such as inhaled corticosteroids (ICS)” ^10^. The meeting report notes that asthma was originally thought to be a disease of airways smooth muscle (i.e. bronchospasm, controlled by bronchodilators) but subsequent guidelines highlighted the role of eosinophilic airways inflammation and anti-inflammatory therapies (inhaled corticosteroids); most patients are still using these same two therapies that were developed more than 30 years ago, and there are no effective alternatives to high-dose oral corticosteroids, which are toxic drugs that are may result in significant and potentially unneccesary morbidity and increased mortality in later life.^10^

Characterisation of asthma inflammatory phenotypes in different settings is therefore important as it will: (i) improve understanding of the aetiological mechanisms of asthma; (ii) identify specific causes; (iii) guide the development of new therapeutic and prevention measures that are effective for all asthmatics in both HICs and LMICs.

The World ASthma Phenotypes (WASP) study is an international collaboration to investigate and characterise asthma phenotypes in HICs and LMICs. The detailed rationale and protocol have been published elsewhere^11^. In this first paper, we present the findings with regards to the main four asthma inflammatory phenotypes^12^: <u>eosinophilic asthma (EA)</u> involving raised eosinophil counts either without (<u>eosinophilic</u>) or with (<u>mixed granulocytic</u>) raised neutrophil counts; <u>non-eosinophilic asthma (NEA)</u> including cases of neutrophilic inflammation of the airways in the absence of eosinophilia (<u>neutrophilic</u>), as well as cases with no apparent inflammation of the airways (<u>paucigranulocyctic).</u> In each centre, we assessed the distribution of these asthma phenotypes, and compared their clinical characteristics, and the phenotype distributions across centres.

## Methods

The detailed study methods have been published elsewhere ^11^, but are briefly summarized here. The study was conducted in five centres; Bristol in the UK (Avon Longitudinal Study of Parents and Children, ALSPAC^13-15^), Wellington in New Zealand, Salvador in Brazil, Quininde in Ecuador and Entebbe in Uganda (Table 1), which were known from the International Study of Asthma and Allergies in Childhood (ISAAC) to have a range of prevalence levels and different environmental exposures^16^.

**Table 1:**
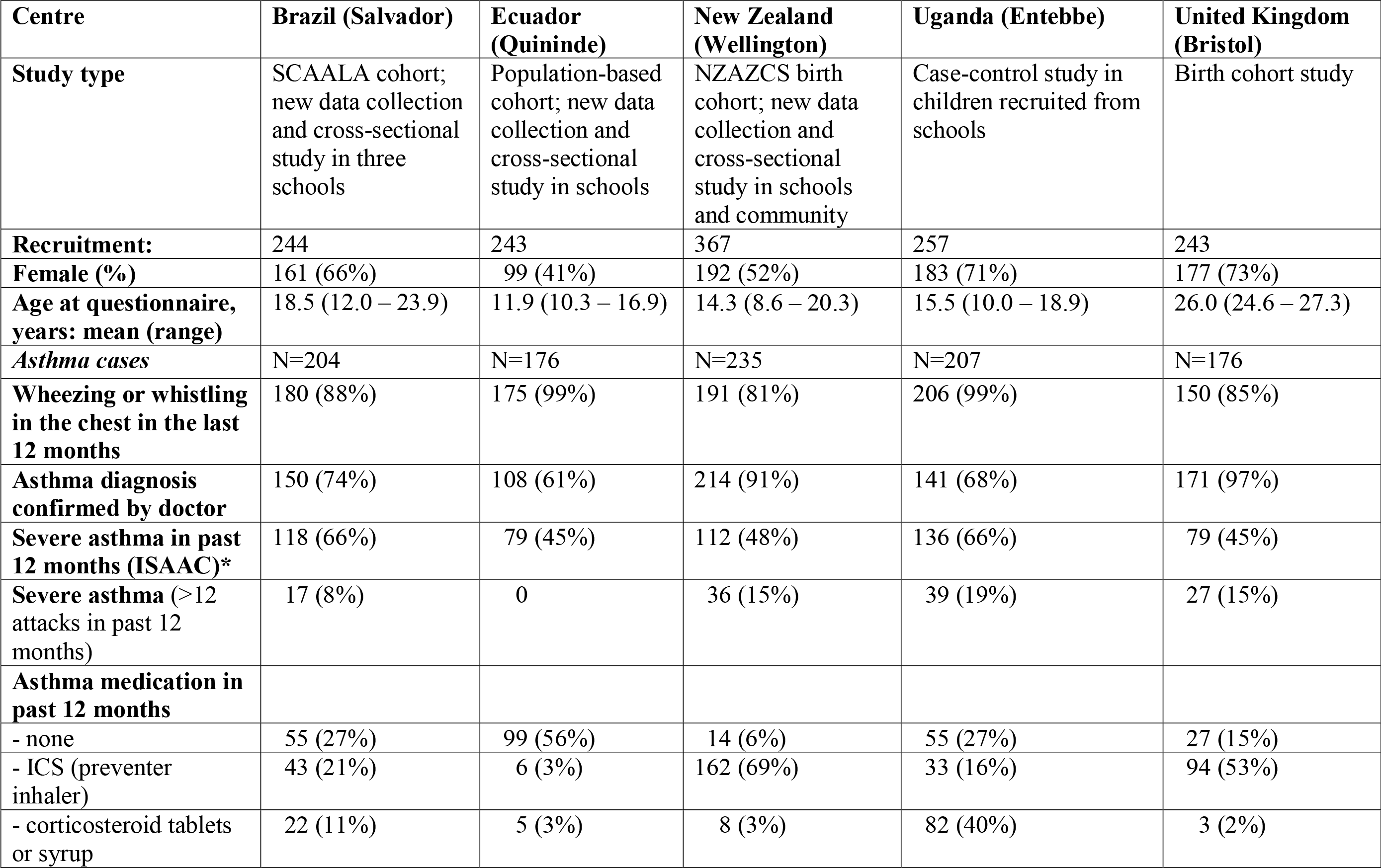
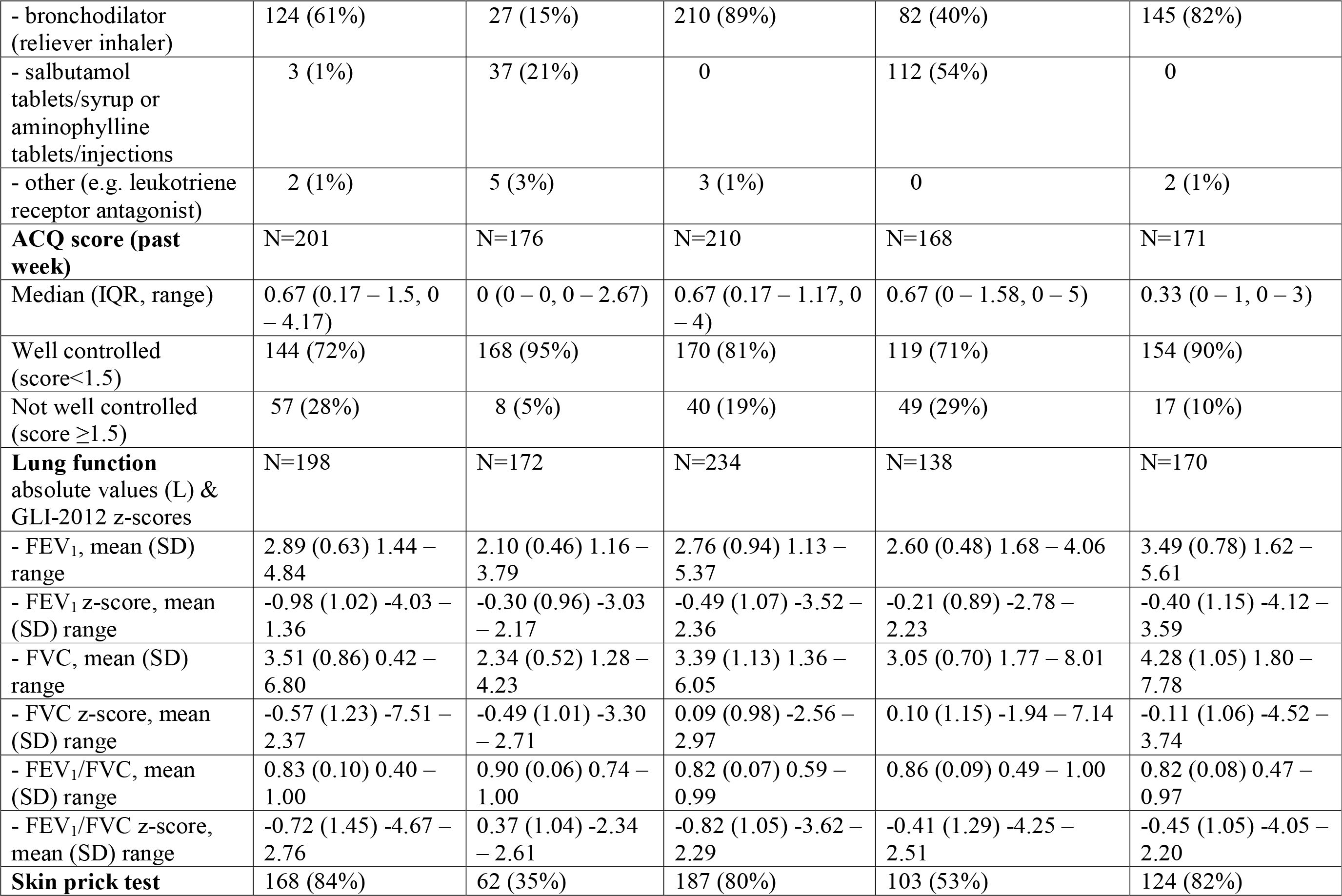
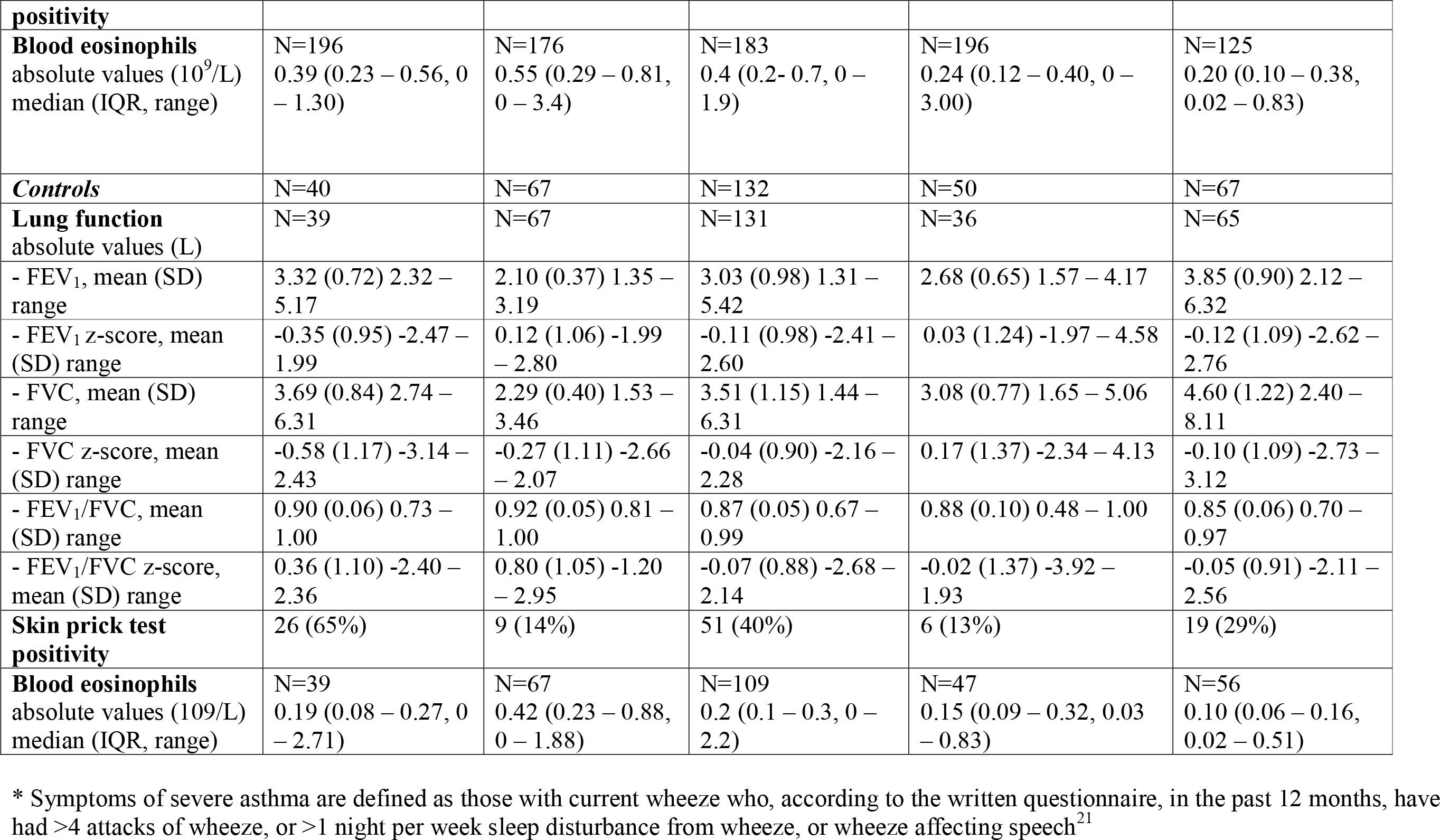
Collaborating studies and centres, and characteristics of study participants.

Ethical approval for the study has been obtained from the LSHTM ethics committee (ref: 9776) and in all five study centres (see Supplementary Information). Informed consent was obtained from all participants or their parents/carers before taking part.

The focus was on defining categories within the groups of asthmatics; however, in each centre, a comparison was also made with non-asthmatics. The recruitment methods differed by centre (Table 1): in four centres (Brazil^17^, Ecuador^17^, New Zealand^18^ and the United Kingdom^13-15^), participants were recruited from ongoing cohort studies, and in three of these centres (Brazil, Ecuador, New Zealand) additional recruitment was made through community recruitment (usually from surveys in schools). In Uganda, participants were recruited from a larger case-control study of asthmatics and non-asthmatics identified through a cross-sectional survey in schools^19^. Asthmatics were identified as those with wheeze or whistling in the chest and/or use of asthma medication in the past 12 months, using the International Study of Asthma and Allergies in Childhood (ISAAC) or (in adults) the European Community Respiratory Health Survey (ECRHS) questionnaires (the same key questions occur in both questionnaires). Non-asthmatics were identified as having no previous or current history of asthma, using the ISAAC and ECRHS questionnaires.

Subjects with chronic disease (except asthma) or who were pregnant were excluded from the study. The study clinic appointment was postponed if participants had a symptomatic respiratory infection in the last 4 weeks or an acute exacerbation of asthma. Participants were asked to stop taking anti-histamines 5 days before the visit, steroid nasal sprays for 7 days before the visit and non-steroidal anti-inflammatories (NSAIDs) for 6 hours before the visit. Asthmatics were asked to stop their asthma medication if safe to do so: no cromoglycate, nedocromil, short acting beta-agonists, ipratropriumbromide for 6 hours prior to visit, no long acting beta-agonists for 12 hours prior to visit, and no theophyllines for 24 hours prior to visit.

### Data collection

Information was collected using standardised methods and operational procedures. These included risk factor questionnaires, clinical characterisation, and blood, and induced sputum. The first 50 asthmatics to be recruited in each center were invited to repeat induced sputum testing after approximately three months

#### Questionnaire

we included questions on the frequency of symptoms, and on use of asthma medications. These were based on the ISAAC Phase II (asthma management) with additional questions on asthma control^20^. We used the standard ISAAC definition of chronic severe asthma as those with current wheeze who have more than 4 attacks of wheezing in the last 12 months, or more than one night per week sleep disturbance, or wheeze affecting speech^21^. In addition, we used a stricter definition of severe asthma of more than 12 attacks of wheezing in the last 12 months, which aligns more closely with the GINA 2008 guidelines.

#### Skin prick tests

Skin prick tests (SPT) were carried out as described previously^22, 23^, with atopy defined as the presence of ≥1 weal of ≥3mm to at least one of a panel of ≥8 commercially available allergens (ALK, Stallergenes, Greer, Immunotech), including house dust mite (*Dermatophagoides pteronyssinus*), tree pollen mix, grass pollen mix, cat and dog dander, *Alternaria tenuis*, Penicillium mix, plus locally relevant allergens^11^ (e.g. Blomia tropicalis (dust mite), German cockroach, American cockroach, Aspergillus fumigatus, Cladosporium). Histamine and saline were used as positive and negative controls, respectively.

#### Lung function testing

Lung function testing was conducted according to American Thoracic Society (ATS) criteria.The Global Lung Function Initiative (GLI-2012) reference values were used to calculate z-scores, taking into account age, sex, height and ethnicity.^24^

#### Exhaled nitric oxide (FeNO)

FeNO was measured in three centres (it was an optional part of the study protocol). In Ecuador and Uganda the Bedfont NOBreath device (Bedfont Scientific Ltd, Maidstone, UK) was used and in New Zealand they used the Hypair FeNO analyser (Medisoft, Sorinnes, Belgium). The Bedfont NOBreath and the Hypair were compared in a subgroup of participants in New Zealand and no substantial differences were observed.

#### Blood eosinophils

15mls of peripheral blood was collected from each participant into two 5ml EDTA tubes. Blood samples were processed within 4 hours. A full blood count was conducted according to standard procedures and included an eosinophil count.

#### Sputum induction

Sputum induction was conducted using a standardised protocol that we have used previously^25^, adapted from the protocol developed by Gibson et al^26^. Aerosolised hypertonic saline (4.5%w/v) was produced using an ultrasonic nebuliser (DeVilbiss Ultraneb 3000, Langen, Germany) and administered orally through a mouthpiece (Hans-Rudolph Inc, Kansas City, USA) for increasing intervals from 30 seconds to 4 minutes, to a total of 15.5 minutes. Spirometry was conducted between intervals to monitor FEV_1_, and salbutamol was administered if FEV_1_ dropped to 75%-predicted or less. Participants were subsequently encouraged to produce sputum in a sterile plastic container. Sputum was processed according to a well□characterized protocol,^27^ and the resulting cell suspension used to prepare cytospin slides stained using a Diff-Quik® fixative and stain set (Dade Behring, Deerfield, IL). The sputum slides were all read in Wellington, New Zealand, with the exception of the Brazil slides, which could not be shipped overseas because of restrictions on samples containing DNA: these were therefore read in Brazil, with a sample of slides being checked (using microscopy images) by the group in Wellington. The asthma inflammatory phenotypes were defined as: <u>eosinophilic</u>: ≥2.5% eosinophils and <61% <u>neutrophils</u>; mixed granulocytic: ≥2.5% eosinophils and ≥61% neutrophils; neutrophilic: <2.5% eosinophils and ≥61% neutrophils; <u>paucigranulocytic:</u> <2.5% eosinophils and <61% neutrophils. We repeated the analysis using a 1% cut-off for eosinophils^12^, and separately using a 54% cut-off for neutrophils as this has been used in other paediatric studies^28, 29^. The results are also presented excluding the low quality slides (those with less than 400 total non-squamous cells, and >30% squamous cells).

#### Data analysis

most analyses involved simple means and percentages, but we calculated 95% confidence intervals (95%CI) for the key means and percentages, as well as odds ratios^30^ and population attributable risks, and 95% CIs where appropriate.

## Results

In Brazil, 685 individuals were contacted (or contact attempted) and 298 (44%) agreed to schedule a clinic visit, of whom 250 attended. Participants were recruited from the SCAALA cohort (14 asthmatics and 15 non-asthmatics) and from schools (190 asthmatics and 25 non-asthmatics). In Ecuador, the response rate was over 95%; participants were recruited from a cohort study (89 asthmatics and 67 non-asthmatics) and from schools (87 asthmatics). In New Zealand, participants were recruited from a larger study. From a birth cohort, 908 individuals were invited and 386 (43%) agreed and completed the questionnaire. Contact details for 1279 individuals were obtained by community recruitment, of whom 824 (64%) confirmed that they wanted to participate (n=455 refused or were ineligible/uncontactable after initial contact). For this study, 134 participants were recruited from the birth cohort (57 asthmatics and 77 non-asthmatics) and 233 from the community (178 asthmatics and 55 non-asthmatics). In Uganda, recruitment was through schools for a larger study; of 6353 parents invited, 1750 (28%) attended a parents’ meeting about the study. Of those attending the meeting, 1734 (99%) consented to their child taking part. 207 asthmatics and 50 non-asthmatics were selected for this study. In the UK, 963 ALSPAC participants were invited; 54 (6%) declined and 390 (40%) accepted; following screening for eligibility, clinic visits were completed for 243.

Table 1 summarizes the characteristics of the participants (the characteristics of the study centres are described in the previously published protocol paper^11^). Overall, we recruited 998 asthma cases and 356 controls. All centres included children and adolescents (age-range 8-20 years), except for the UK centre for which the participants were 26-27 years. The proportion of cases who were skin-prick-test (SPT) positive ranged from 35% (Ecuador) to 84% (Brazil). The proportion of controls who were SPT+ ranged from 13% (Uganda) to 65% (Brazil). We also calculated the population attributable risks of skin-prick-test positivity for asthma (not shown in table): Brazil (50%), Ecuador (25%), New Zealand (67%), Uganda (43%) and United Kingdom (59%), with an overall estimate of 48% (95%CI 44%-52%%).

The proportion of participants who produced a sputum sample ranged from 74% (Brazil) to 93% (UK) (Table 2). Of these, the proportion of countable sputum slides ranged from 46% (UK) to 95% (New Zealand). The proportion with eosinophilic asthma (EA), i.e. either eosinophilic or mixed granulocytic inflammatory cell types was one-half (50%) in New Zealand, 33% in the UK, and about one-third (32%-35%) in the LMIC centres. Regarding the inflammatory subtypes, the findings were reasonably consistent across centres, with a predominance of eosinophilic and paucigranulocytic asthma, neither of which involve high levels of neutrophils. The exception was Uganda, where a high proportion of non-eosinophilic cases had elevated levels of neutrophils. However, this proportion was higher in the controls (60%) than in the cases (35%). We also examined the repeatability of the phenotyping with a repeat sputum induction test done 3-6 months after the original test. Overall, 68% (95%CI 61%-74%) had the same general phenotype (EA or NEA) across the two tests, with roughly equal numbers switching from EA to NEA and vice versa.

**Table 2:** Sputum slide results by centre.

| **Centre** | **Brazil** | **Ecuador** | **New Zealand** | **Uganda** | **United Kingdom** | **Total** |
| --- | --- | --- | --- | --- | --- | --- |
| Number (%) of participants who provided sputum sample | 181 (74%) | 244 (91%) | 350 (88%) | 221 (86%) | 229 (93%) | 1225 (87%) |
| Number of participants with countable sputum slide(s) (% of those who provided sample) | 137 (76%) | 183 (75%) | 332 (95%) | 117 (53%) | 106 (46%) | 875 (71%) |
| Sputum inflammatory phenotype: **asthma cases** | N=115 | N=125 | N=207 | N=98 | N=76 | N=621 |
| eosinophilic | 38 (33%) | 35 (28%) | 99 (48%) | 25 (25%) | 23 (30%) | 220 (35%) |
| mixed granulocytic | 2 (2%) | 5 (4%) | 5 (2%) | 8 (8%) | 2 (3%) | 22 (4%) |
| neutrophilic | 5 (4%) | 8 (6%) | 14 (7%) | 34 (35%) | 6 (8%) | 67 (11%) |
| paucigranulocytic | 70 (61%) | 77 (62%) | 89 (43%) | 31 (32%) | 45 (59%) | 312 (50%) |
| **Repeat sputum slide** |  |  |  |  |  |  |
| same phenotype (EA or NEA*) | 27 (68%) | 25 (69%) | 72 (67%) | 9 (75%) | 5 (56%) | 138 (68%) |
| Changed: |  |  |  |  |  |  |
| EA to NEA | 6 | 4 | 18 |  | 3 | 31 (15%) |
| NEA to EA | 7 | 7 | 17 | 3 | 1 | 35 (17%) |
| Sputum inflammatory phenotype: **controls** | N=20 | N=41 | N=104 | N=20 | N=29 | N=214 |
| eosinophilic | 4 (20%) | 3 (7%) | 11 (11%) | 2 (10%) | 3 (10%) | 23 (11%) |
| mixed granulocytic | 0 | 0 | 1 (1%) | 1 (5%) | 0 | 2 (1%) |
| neutrophilic | 4 (20%) | 1 (2%) | 11 (11%) | 12 (60%) | 3 (10%) | 31 (14%) |
| paucigranulocytic | 12 (60%) | 37 (90%) | 81 (78%) | 5 (25%) | 23 (79%) | 158 (74%) |

\* EA (eosinophilic or mixed); NEA (neutrophilic or paucigranulocytic)

The characteristics of the four inflammatory phenotypes in each centre are summarized in Table 3, and are given in more detail in Appendix 1. Using the ISAAC definition^21^ of asthma severeity in the last 12 months severe asthma was more common in the eosinophilic than in the non-eosinophilic phenotypes (p<0.001): the proportions with severe asthma were 60% for eosinophilic, 68% for mixed granulocytic, 43% for neutrophilic, and 43% for paucigranulocytic. The corresponding proportions with well-controlled asthma in the past week were 78%, 64%, 81%, and 79% respectively. The mean FEV1 scores tended to be lower in the EA (eosinophilic and mixed granulocytic) groups than in the NEA (neutrophilic and paucigranulocytic) groups. We also calculated the population attributable risks of EA (eosinophilic or mixed granulocytic) for asthma (not shown in table): Brazil (18%), Ecuador (27%), New Zealand (44%), Uganda (22%) and United Kingdom (18%), with an overall estimate of 30% (95%CI 25%-33%).

**Table 3:**
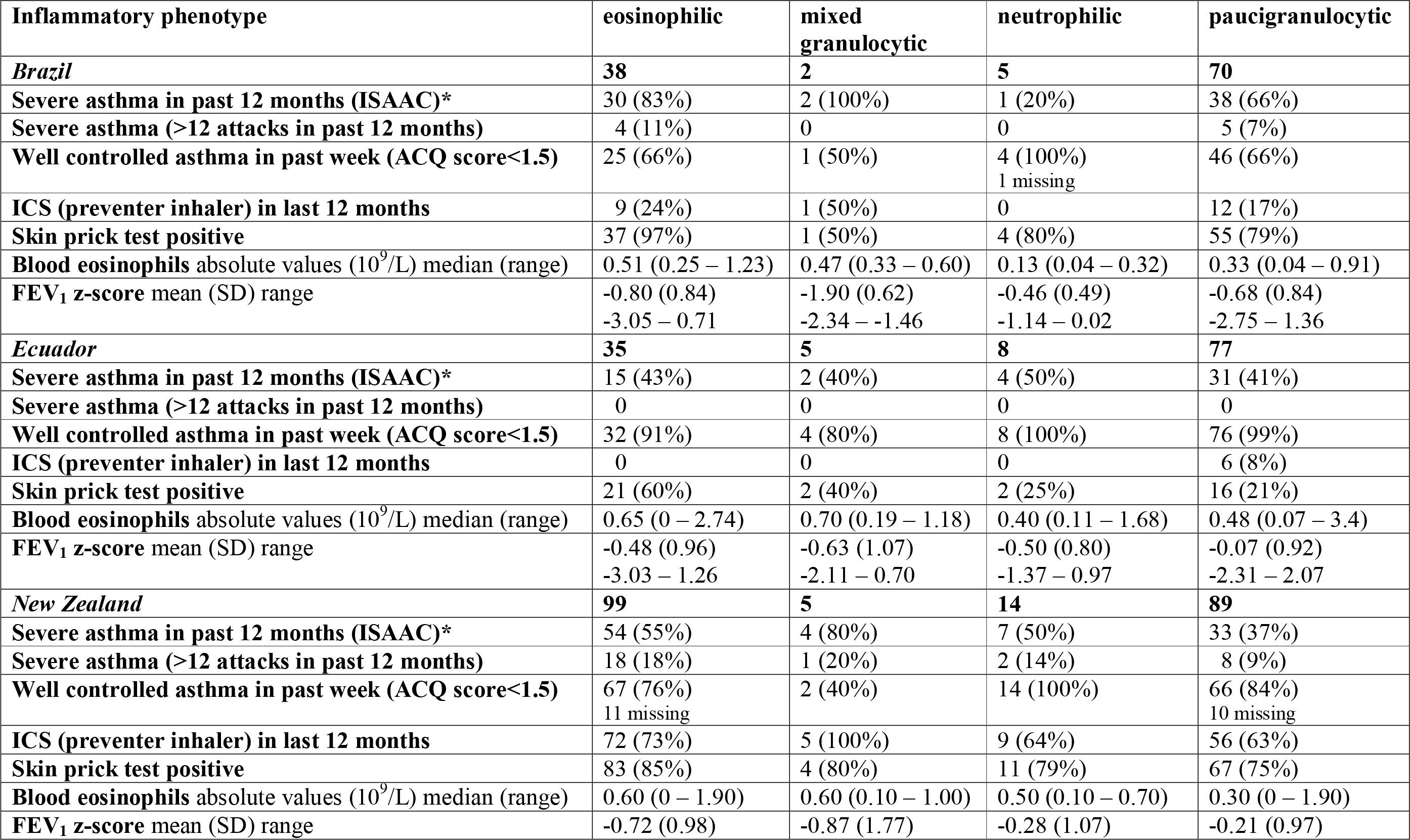
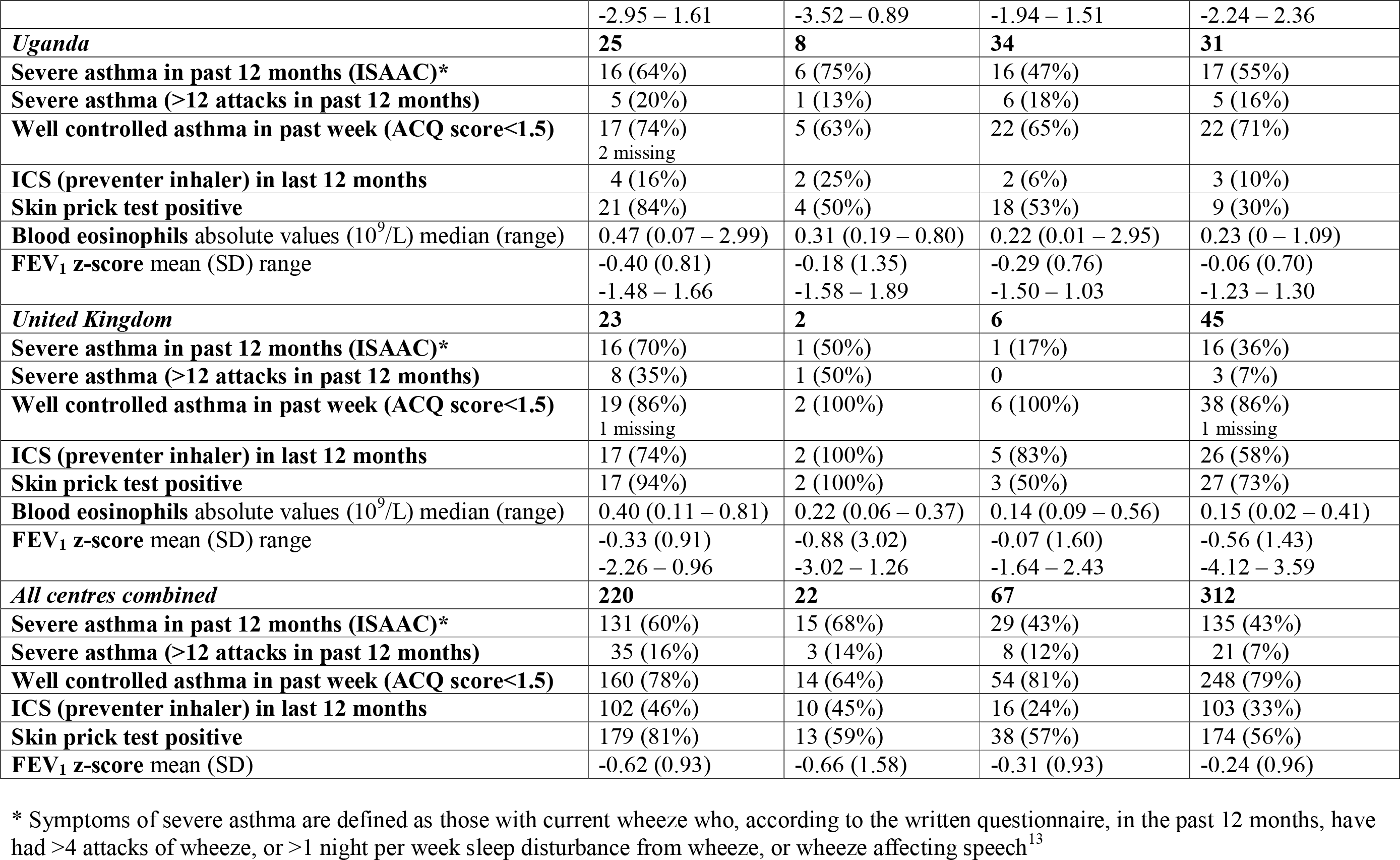
Characteristics by inflammatory phenotype in each centre.

Appendix 2 shows the sputum results excluding low quality slides (those with less than 400 total non-squamous cells, and ≥30% squamous cells). The findings changed little; for example, the proportions with eosinophilic or mixed granulocytic asthma were unchanged (33%) in the UK, and changed from 50% to 52% in New Zealand, 35% to 32% in Brazil, 32% to 33% in Ecuador, and 33% to 29% in Uganda. Using a 1% eosinophil cut-off (instead of 2.5%), the proportions with EA ranged from 40% (Ecuador) to 69% (New Zealand) (Appendix 3). Changing the neutrophil cut-off to 54% (instead of 61%) did not alter the findings very much in most centres; the largest changes in the proportion who were neutrophilic were 35% to 40% in Uganda and 8% to 14% in the UK (Appendix 3).

## Discussion

This is the first time that sputum induction has been used in a standardised manner to compare asthma inflammatory phenotypes in centres in high income countries (HICs) and low-and-middle-income countries (LMICs). The participants were chosen to be a representative sample of asthmatics in general, rather than focussing on severe asthma, and this is reflected in the clinical indicators. Nevertheless, using the ISAAC definition^21^ (which is based on symptoms in the previous year), the proportion of participants with severe asthma over the previous 12 months (ISAAC definition) ranged from 45% (UK, Ecuador) to 66% (Brazil, Uganda). The proportions with well-controlled asthma in the past week ranged from 71% (Uganda) to 95% (Ecuador).

There are several key findings that should be considered.

Firstly, for the four centres which involved children and adolescents, this study confirms previous research in HICs that only about one-half, or less, of current asthmatics have EA, and it shows for the first time that only about one-third of asthmatics in the centres in LMICs have EA. This adds to previous evidence that there is a substantial proportion of asthma cases that involve non-eosinophilic inflammatory phenotypes, and shows for the first time to our knowledge, that this is the case in asthmatics in LMICs as well as HICs. Some, but not all of these NEA cases may represent cases of EA which have responded to corticosteroid treatment, but it appears unlikely that this accounts for all of the NEA case^12^s. Furthermore, even if their sputum profiles can in part be explained by treated EA, most have persistent symptoms. The proportions of asthma cases who were classified as eosinophilic or mixed granulocytic asthma (the two inflammatory phenotypes which involve eosinophilia, based on >2.5% of cells being eosinophils) were 32%-35% in the LMICs (Brazil, Ecuador, Uganda); in the HICs it was 50% in New Zealand, and 33% in the United Kingdom. It is possible that this difference between the United Kingdom and New Zealand is due to the different age-groups that were studied. However, it is reasonably consistent with the findings of the ISAAC Phase II study^5^ which was conducted in 8-12 year-olds: the overall proportions with positive skin prick tests were 35.6% in New Zealand compared with 17.5% in the UK; the proportion of asthma cases (wheeze in the last year) that were skin-prick-test positive were 60% and 39% respectively. Thus, it is possible that this finding represents a real difference between the United Kingdom and New Zealand, which warrants further investigation, given that both are high-income countries with similar levels of asthma prevalence^16^.

Secondly, there was a high proportion of neutrophilic cases in Uganda. The high proportion in the cases may be in part due to the low proportion with eosinophilia, but the proportion was actually lower in the asthma cases (35%) than in the controls (50%). Previous studies have found neutrophilia to be associated with more severe asthma^12^, but this was not observed in the Uganda cases in the current study (Table 3). Thus it is possible that the high proportion with neutrophilia reflects non-asthmatic neutrophilic inflammation due to environmental exposures (e.g. indoor air pollution, increased risk of infections, exposure to animals),. It is notable, however, that all centres (with the exception of Ecuador, with small numbers) actually showed a higher proportion of neutrophilia in the controls (14% overall) than in the cases (11%). It is possible therefore that some of the neutrophilic cases have neutrophilia incidentally and it is not integral to their asthma.

Thirdly, the most striking finding is the high proportion of cases with neither eosinophilia or neutrophilia (i.e. paucigranulocytic asthma). This confirms findings from previous studies in HICs^25^, which have shown that a high proportion of asthmatics appear to have no airways inflammation, thus raising the possibility that non-inflammatory mechanisms (e.g. neural mechanisms^31^) may be involved.

Two objections may be raised against this hypothesis. One is that the inflammatory phenotype is unstable, and that children with inherently eosinophilic asthma may only show eosinophilic inflammation from time to time, particularly when they are having attacks. However, previous studies have shown the non-inflammatory phenotype to be relatively stable^12, 32, 33^, and we have found similar findings here (although with a slightly lower level of reproducibility): 68% had the same phenotype (EA or NEA) in the repeat sputum assessments. A second objection might be that those with paucigranulocytic asthma may not really have asthma, or may have extremely mild asthma; alternatively, it could represent low-level eosinophilic inflammation occurring outside the central airways^34^. We found only small differences in chronic asthma severity between eosinophilic (60%), mixed granulocytic (68%), neutrophilic (43%) and paucigranulocytic (43%) asthma. Although the definition of chronic severity that we used (based on the ISAAC definition) produces relatively high severity estimates, we used a consistent definition across centres and inflammatory phenotypes, so it is noteworthy that we did not see major differences in severity between the phenotypes. These proportions are relatively high, given that most participants had well-controlled asthma, but this reflects the ISAAC definition which is based on symptoms in the last year and yields higher estimates of chronic asthma severity than do other definitions which focus on acute clinical severity. On the other hand, most participants had well-controlled asthma in the previous week (Table 3).

Some of the limitations of this study should also be acknowledged.

We endeavoured to obtain random population samples of asthmatics in each centre, by taking random samples in schools, and by utilizing existing cohort studies. Standardizing the data collection (particularly the sputum induction) was difficult, and not all centres could obtain readable sputum samples from a high proportion of participants. There were also difficulties in reading some of the slides (particularly those with high proportions of squamous cells). In particular, only 46% of the slides from Bristol were readable (table 2), even though a high proportion of participants (93%) provided samples. The reasons for this are unclear, but the findings did not change markedly when we restricted our analyses to high quality slides (Appendix 2).

In conclusion, this study confirms that most cases are non-eosinophilic with most having no sign of airways inflammation, with the proportions ranging from 50% to 65%, across varied environments. This has potentially major implications for asthma prevention and management in both of these contexts (HICs and LMICs), and supports the urgent need to recognise that asthma is a heterogeneous condition, and to develop new therapies which target NEA^9^. In particular, currently we are treating a large proportion of asthmatics with drugs that are likely to be ineffective and potentially harmful. In addition, the causal environmental exposures and triggers are likely to be different for EA and NEA. However, due to a lack of research into NEA, we know very little about environmental (or other) causes of NEA, hampering the development of more effective interventions. In addition, the development of more effective treatment options for NEA requires urgent attention.

## Data Availability

Data is currently not available. We will consider requests for data sharing after the main findings have been published.

## Acknowledgements

The World ASthma Phenotypes (WASP) collaboration is based on the AsthmaPhenotypes study which is funded by the European Research Council under the European Union’s Seventh Framework Programme (FP7/2007-2013) / ERC grant agreement no 668954.

ALSPAC: We are extremely grateful to all the families who took part in this study, the midwives for their help in recruiting them, and the whole ALSPAC team, which includes interviewers, computer and laboratory technicians, clerical workers, research scientists, volunteers, managers, receptionists and nurses. The UK Medical Research Council and Wellcome (Grant ref: 21706/Z/19/Z) and the University of Bristol provide core support for ALSPAC. This publication is the work of the authors and Lucy Pembrey and Neil Pearce will serve as guarantors for the contents of this paper. A comprehensive list of grants funding is available on the ALSPAC website (http://www.bristol.ac.uk/alspac/external/documents/grant-acknowledgements.pdf); this research was specifically funded by the ERC grant agreement no 668954 (see above). Alison Elliott and Philip Cooper were funded by Wellcome Trust grant numbers 095778 and 088862, respectively. We would like to thank the participants of this study in all five WASP centres.

## The WASP Study group

<u>United Kingdom, London:</u> Neil Pearce*, Lucy Pembrey*, Steven Robertson, Karin van Veldhoven, Sinead Langan, Sarah Thorne, Donna Davoren

<u>United Kingdom, Bristol:</u> John Henderson, Susan Ring, Elizabeth Brierley, Sophie Fitzgibbon, Simon Scoltock, Amanda Hill

<u>Brazil, Leading Group:</u> Alvaro Cruz*, Camila Figueiredo*, Mauricio Barreto*

<u>Brazil, ProAR collaborators/associates:</u> Cinthia Vila Nova Santana, Gabriela Pimentel, Gilvaneide Lima, Valmar Bião Lima, Jamille Fernandes

<u>Brazil, Lab students/associates:</u> Tamires Cana Brasil Carneiro, Candace Andrade, Gerson Queiroz, Anaque Pires, Milca Silva, Jéssica Cerqueira

<u>Ecuador:</u> Phil Cooper*, Martha Chico, Cristina Ardura-Garcia, Araceli Falcones, Aida Y Orviedo, Andrea Zambrano

<u>New Zealand:</u> Jeroen Douwes*, Collin Brooks*, Hajar Ali, Jeroen Burmanje

<u>Uganda:</u> Harriet Mpairwe*, Irene Nambuya, Pius Tumwesige, Milly Namutebi, Marble Nnaluwooza, Mike Mukasa

*Writing Group for this paper

## References

1. Edwards, M.R., et al., Addressing unmet needs in understanding asthma mechanisms: From the European Asthma Research and Innovation Partnership (EARIP) Work Package (WP)2 collaborators. Eur Respir J, 2017. 49(5).

2. Pavord, I.D., et al., After asthma: redefining airways diseases. Lancet, 2017.

3. Douwes, J., et al., Non-eosinophilic asthma: importance and possible mechanisms.[comment]. Thorax., 2002. 57(7): p. 643–8.

4. Pearce, N., J. Pekkanen, and R. Beasley, How much asthma is really attributable to atopy? Thorax., 1999. 54(3): p. 268–72.

5. Weinmayr, G., et al., Atopic sensitization and the international variation of asthma symptom prevalence in children. American Journal of Respiratory and Critical Care Medicine, 2007. 176(6): p. 565–574.

6. Barreto, M.L., et al., Poverty, dirt, infections and non-atopic wheezing in children from a Brazilian urban center. Respiratory Research, 2010. 11.

7. Strina, A., et al., Risk factors for non-atopic asthma/wheeze in children and adolescents: a systematic review. Emerg Themes Epidemiol, 2014. 11: p. 5.

8. Brown, H.M., Treatment of chronic asthma iwht predniosolone: significance of eosinophils in the sputum. Lancet, 1958. 2: p. 1245–1247.

9. Pavord, I.D., et al., After asthma: redefining airways diseases. Lancet, 2018. 391(10118): p. 350–400.

10. Asthma still kills: Urgent priorities for the international research community to treat, prevent and cure asthma. 2019, Asthma UK: London.

11. Pembrey, L., et al., Understanding asthma phenotypes: the World Asthma Phenotypes (WASP) international collaboration. ERJ Open Research, 2018. 4: p. 00013–2018.

12. Simpson, J.L., et al., Inflammatory subtypes in asthma: Assessment and identification using induced sputum. Respirology, 2006. 11(1): p. 54–61.

13. Boyd, A., et al., Cohort Profile: the ‘children of the 90s’--the index offspring of the Avon Longitudinal Study of Parents and Children. Int J Epidemiol, 2013. 42(1): p. 111–27.

14. Fraser, A., et al., Cohort Profile: the Avon Longitudinal Study of Parents and Children: ALSPAC mothers cohort. Int J Epidemiol, 2013. 42(1): p. 97–110.

15. Northstone, K., et al., The Avon Longitudinal Study of Parents and Children (ALSPAC): an update on the enrolled sample of index children in 2019. Wellcome Open Res, 2019. 4: p. 51.

16. Asher, M.I., et al., Worldwide variations in the prevalence of asthma symptoms: International Study of Asthma and Allergies in Childhood (ISAAC). European Respiratory Journal, 1998. 12: p. 315–335.

17. Barreto, M.L., et al., Risk factors and immunological pathways for asthma and other allergic diseases in children: background and methodology of a longitudinal study in a large urban center in Northeastern Brazil (Salvador-SCAALA study). BMC Pulm Med, 2006. 6: p. 15.

18. Epton, M.J., et al., The New Zealand Asthma and Allergy Cohort Study (NZA2CS): assembly, demographics and investigations. BMC Public Health, 2007. 7: p. 26.

19. Mpairwe, H., et al., Risk factors for asthma among schoolchildren who participated in a case-control study in urban Uganda. Elife, 2019. 8.

20. Juniper, E.F., et al., Asthma Control Questionnaire in children: validation, measurement properties, interpretation. Eur Respir J, 2010. 36(6): p. 1410–6.

21. Lai, C.K., et al., Global variation in the prevalence and severity of asthma symptoms: phase three of the International Study of Asthma and Allergies in Childhood (ISAAC). Thorax, 2009. 64(6): p. 476–83.

22. Wickens, K., et al., Farm residence and exposures and the risk of allergic diseases in New Zealand children. Allergy, 2002. 57(12): p. 1171–9.

23. Weiland, S.K., et al., Phase II of the international study of asthma and allergies in childhood (ISAAC II): rationale and methods. European Respiratory Journal, 2004. 24(3): p. 406–412.

24. Quanjer, P.H., et al., Multi-ethnic reference values for spirometry for the 3-95-yr age range: the global lung function 2012 equations. Eur Respir J, 2012. 40(6): p. 1324–43.

25. Brooks, C.R., et al., Absence of airway inflammation in a large proportion of adolescents with asthma. Respirology, 2016. 21(3): p. 460–466.

26. Gibson, P.G., et al., Epidemiological association of airway inflammation with asthma symptoms and airway hyperresponsiveness in childhood. Am J Respir Crit Care Med, 1998. 158(1): p. 36–41.

27. Gibson, P.G., Use of induced sputum to examine airway inflammation in childhood asthma. J Allergy Clin Immunol, 1998. 102(5): p. S100–1.

28. Eller, M.C.N., et al., Can inflammatory markers in induced sputum be used to detect phenotypes and endotypes of pediatric severe therapy-resistant asthma? Pediatr Pulmonol, 2018. 53(9): p. 1208–1217.

29. Fleming, L., et al., Sputum inflammatory phenotypes are not stable in children with asthma. Thorax, 2012. 67(8): p. 675–81.

30. Pearce, N., Effect measures in prevalence studies. Environmental Health Perspectives, 2004. 112(10): p. 1047–1050.

31. Baroffio, M., et al., Noninflammatory mechanisms of airway hyper-responsiveness in bronchial asthma: an overview. Ther Adv Respir Dis, 2009. 3(4): p. 163–74.

32. Simpson, J.L., P. McElduff, and P.G. Gibson, Assessment and reproducibility of non-eosinophilic asthma using induced sputum. Respiration, 2010. 79(2): p. 147–51.

33. van Veen, I.H., et al., Consistency of sputum eosinophilia in difficult-to-treat asthma: a 5-year follow-up study. J Allergy Clin Immunol, 2009. 124(3): p. 615-7, 617.e1-2.

34. Demarche, S., et al., Detailed analysis of sputum and systemic inflammation in asthma phenotypes: are paucigranulocytic asthmatics really non-inflammatory? BMC Pulm Med, 2016. 16: p. 46.

