## Supplementary Appendix for "Asthma inflammatory phenotypes on four continents: most asthma cases are non-eosinophilic"

**Online appendix 1: Centre-specific participant and clinical characteristics by inflammatory phenotype**

**Brazil**

| **Inflammatory phenotype** | **eosinophilic** | **mixed granulocytic** | **neutrophilic** | **paucigranulocytic** | Asthmatics without sputum result | **Controls** (with sputum results) |
| --- | --- | --- | --- | --- | --- | --- |
|  | **38** | **2** | **5** | **70** | **89** | **20** |
| Female (%) | 24 (63%) | 2 (100%) | 4 (80%) | 51 (73%) | 53 (60%) | 15 (75%) |
| Age at questionnaire, years: mean (range) | 17.8 (12.5 – 23.8) | 18.3 (18.0 – 18.6) | 17.6 (16.3 – 19.0) | 18.4 (13.1 – 23.1) | 18.4 (12.0 – 23.9) | 19.7 (17.3 – 23.0) |
| Asthma diagnosis confirmed by doctor | 30 (79%) | 2 (100%) | 3 (60%) | 44 (63%) | 71 (80%) | - |
| Age at asthma diagnosis, years: median (range) | 7 (1 – 23) | 5 ( 5 – 5) | 2 (1 – 5) | 7 (1 – 20) | 5 (0 – 18) | - |
| missing | 11 | 1 | 2 | 29 | 26 |  |
| **Asthma severity in past 12 months*** |  |  |  |  |  | - |
| mild or moderate | 6 (17%) | 0 | 4 (80%) | 20 (34%) | 32 (41%) |  |
| severe | 30 (83%) | 2 (100%) | 1 (20%) | 38 (66%) | 47 (59%) |  |
| missing | 2 | 0 | 0 | 12 | 10 |  |
| **Severe asthma** (>12 attacks in past 12 months) | 4 (11%) | 0 | 0 | 5 (7%) | 8 (9%) |  |
| **Asthma medication in past 12 months** |  |  |  |  |  | - |
| none | 8 (21%) | 0 | 3 (60%) | 23 (33%) | 21 (24%) |  |
| ICS (preventer inhaler) | 9 (24%) | 1 (50%) | 0 | 12 (17%) | 21 (24%) |  |
| Bronchodilator (reliever inhaler) | 22 (58%) | 1 (50%) | 2 (40%) | 42 (60%) | 57 (64%) |  |
| **ACQ score (past week)** |  |  |  |  |  | - |
| Median (IQR, range) | 0.92 (0.17 – 1.83, 0 – 2.83) | 1.83 (0.33 – 3.33, 0.33 – 3.33) | 0.17 (0.08 – 0.33, 0 – 0.5) | 1.0 (0.33 – 1.67, 0 – 3) | 0.5 (0.17 – 1.33, 0 – 4.17) |  |
| Well controlled (score<1.5) | 25 (66%) | 1 (50%) | 4 (100%) | 46 (66%) | 68 (78%) |  |
| Not well controlled (score ≥1.5) | 13 (34%) | 1 (50%) | 0 | 24 (34%) | 19 (22%) |  |
| missing | 0 | 0 | 1 | 0 | 2 |  |
| **Lung function** absolute values (L) & GLI-2012 z-scores | n=37 | n=2 | n=5 | n=68 | n=86 | n=19 |
| - FEV_1_, mean (SD) range | 2.98 (0.66)  1.97 – 4.48 | 2.23 (0.11)  2.15 – 2.31 | 3.20 (0.74)  2.44 – 4.33 | 2.92 (0.48)  1.64 – 3.99 | 2.84 (0.70)  1.44 – 4.84 | 3.35 (0.73)  2.50 – 4.73 |
| - FEV_1_ z-score, mean (SD) range | -0.80 (0.84)  -3.05 – 0.71 | -1.90 (0.62)  -2.34 – -1.46 | -0.46 (0.49)  -1.14 – 0.02 | -0.68 (0.84)  -2.75 – 1.36 | -1.30 (1.16)  -4.03 – 0.90 | -0.21 (0.94)  -1.49 – 1.99 |
| - FVC, mean (SD) range | 3.69 (0.85)  2.20 – 5.99 | 1.81 (1.96)  0.42 – 3.19 | 3.63 (0.78)  2.88 – 4.87 | 3.46 (0.62)  2.43 – 4.80 | 3.50 (0.98)  0.91 – 6.80 | 3.76 (0.80)  2.80 – 5.24 |
| - FVC z-score, mean (SD) range | -0.15 (0.99)  -2.64 – 1.63 | -3.63 (5.49)  -7.51 – 0.25 | -0.41 (0.49)  -1.02 – 0.20 | -0.37 (0.86)  -2.64 – 2.37 | -0.84 (1.32)  -5.95 – 2.01 | -0.33 (1.19)  -2.64 – 2.43 |
| - FEV_1_/FVC, mean (SD) range | 0.81 (0.09)  0.64 – 0.99 | 0.73 (0.13)  0.63 – 0.82 | 0.88 (0.03)  0.85 – 0.92 | 0.85 (0.09)  0.63 – 1.00 | 0.82 (0.12)  0.40 – 1.00 | 0.88 (0.06)  0.73 – 0.99 |
| - FEV_1_/FVC z-score, mean (SD) range | -1.04 (1.29)  -3.14 – 2.18 | -2.25 (1.22)  -3.12 – -1.39 | -0.19 (0.54)  -0.81 – 0.35 | -0.46 (1.28)  -3.18 – 2.23 | -0.79 (1.63)  -4.67 – 2.76 | 0.06 (1.11)  -2.40 – 1..87 |
| **Skin prick test positive** | 37 (97%) | 1 (50%) | 4 (80%) | 55 (79%) | 71 (83%) | 14 (70%) |
| Not done |  |  |  |  | 3 |  |
| **Blood eosinophils** absolute values (10^9^/L) median (range) | n=36  0.51  (0.25 – 1.23) | n=2  0.47  (0.33 – 0.60) | n=5  0.13  (0.04 – 0.32) | n=66  0.33  (0.04 – 0.91) | n=87  0.35  (0.002 – 1.30) | n=20  0.15  (0.04 – 2.71) |

**Ecuador**

| **Inflammatory phenotype** | **eosinophilic** | **mixed granulocytic** | **neutrophilic** | **paucigranulocytic** | Asthmatics without sputum result | **Controls** (with sputum results) |
| --- | --- | --- | --- | --- | --- | --- |
|  | **35** | **5** | **8** | **77** | **51** | **41** |
| Female (%) | 10 (29%) | 2 (40%) | 3 (38%) | 44 (57%) | 17 (33%) | 11 (27%) |
| Age at questionnaire, years: mean (range) | 12.4  (10.5 – 16.2) | 13.8  (11.7 – 16.8) | 12.3  (10.3 – 14.6) | 11.9  (10.3 – 16.9) | 11.8  (10.3 – 14.3) | 11.7  (11.0 – 12.1) |
| Asthma diagnosis confirmed by doctor | 29 (83%) | 5 (100%) | 5 (63%) | 40 (52%) | 29 (57%) | - |
| Age at asthma diagnosis, years: median (range) | 4 (0.08 – 9) | 4 (0.7 – 13) | 1.3 (0.3 – 6) | 2 (0 – 14) | 2 (0 – 10) | - |
| **Asthma severity in past 12 months*** |  |  |  |  |  | - |
| mild or moderate | 20 (57%) | 3 (60%) | 4 (50%) | 45 (59%) | 24 (47%) |  |
| severe | 15 (43%) | 2 (40%) | 4 (50%) | 31 (41%) | 27 (53%) |  |
| missing |  |  |  | 1 |  |  |
| **Severe asthma** (>12 attacks in past 12 months) | 0 | 0 | 0 | 0 | 0 |  |
| **Asthma medication in past 12 months** |  |  |  |  |  | - |
| none | **13 (37%)** | **1 (20%)** | **5 (63%)** | **52 (68%)** | **28 (55%)** |  |
| ICS (preventer inhaler) | 0 | 0 | 0 | **6 (8%)** | 0 |  |
| Bronchodilator (reliever inhaler) | **7 (20%)** | **1 (20%)** | **0** | **10 (13%)** | **9 (18%)** |  |
| **ACQ score (past week)** |  |  |  |  |  |  |
| Median (IQR, range) | 0 (0 – 0, 0 – 2.67) | 0 (0 – 0, 0 – 2) | 0 (0 – 0, 0 – 0) | 0 ( 0 – 0, 0 – 1.5) | 0 (0 – 0, 0 – 2.67) | - |
| Well controlled (score<1.5) | 32 (91%) | 4 (80%) | 8 (100%) | 76 (99%) | 48 (94%) |  |
| Not well controlled (score ≥1.5) | 3 (9%) | 1 (20%) | 0 | 1 (1%) | 3 (6%) |  |
| **Lung function** absolute values (L) & GLI-2012 z-scores | n=34 | n=5 | n=8 | n=77 | n=48 | n=41 |
| - FEV_1_, mean (SD) range | 2.11 (0.56)  1.16 – 3.75 | 2.36 (0.52)  1.62 – 3.00 | 2.03 (0.49)  1.58 – 2.86 | 2.15 (0.43)  1.37 – 3.79 | 2.00 (0.39)  1.31 – 2.88 | 2.05 (0.35)  1.35 – 3.19 |
| - FEV_1_ z-score, mean (SD) range | -0.48 (0.96)  -3.03 – 1.26 | -0.63 (1.07)  -2.11 – 0.70 | -0.50 (0.80)  -1.37 – 0.97 | -0.07 (0.92)  -2.31 – 2.07 | -0.49 (1.01)  -2.46 – 2.17 | -0.002 (0.93)  -1.99 – 2.80 |
| - FVC, mean (SD) range | 2.39 (0.65)  1.28 – 4.21 | 2.58 (0.52)  1.76 – 3.07 | 2.25 (0.61)  1.69 – 3.38 | 2.36 (0.46)  1.49 – 4.23 | 2.25 (0.48)  1.34 – 3.29 | 2.25 (0.38)  1.57 – 3.46 |
| - FVC z-score, mean (SD) range | -0.56 (0.95)  -3.21 – 1.45 | -0.86 (1.29)  -2.25 – 0.45 | -0.69 (1.10)  -1.78 – 1.01 | -0.33 (0.94)  -2.75 – 1.53 | -0.62 (1.11)  -3.30 – 2.71 | -0.34 (0.94)  -2.25 – 2.07 |
| - FEV_1_/FVC, mean (SD) range | 0.89 (0.06)  0.75 – 1.00 | 0.91 (0.06)  0.85 – 1.00 | 0.91 (0.03)  0.85 – 0.93 | 0.91 (0.05)  0.74 – 1.00 | 0.89 (0.06)  0.77 – 0.99 | 0.91 (0.05)  0.81 – 0.99 |
| - FEV_1_/FVC z-score, mean (SD) range | 0.16 (1.07)  -1.91 – 2.29 | 0.38 (1.22)  -0.87 – 2.29 | 0.35 (0.68)  -0.99 – 1.19 | 0.54 (1.01)  -2.34 – 2.61 | 0.26 (1.11)  -1.76 – 2.47 | 0.67 (0.96)  -1.20 – 2.46 |
| **FeNO level** |  |  |  |  |  |  |
| normal | 12 | 3 | 8 | 60 | 29 | 32 |
| elevated | 23 (66%) | 2 (40%) | 0 (0%) | 17 (22%) | 21 (42%) | 9 (22%) |
| not measured |  |  |  |  | 1 |  |
| **Skin prick test positive** | 21 (60%) | 2 (40%) | 2 (25%) | 16 (21%) | 21 (41%) | 7 (17%) |
| **Blood eosinophils** absolute values (10^9^/L) median (range) | n=35  0.65  (0.002 – 2.74) | n=5  0.70  (0.19 – 1.18) | n=8  0.40  (0.11 – 1.68) | n=77  0.48  (0.07 – 3.40) | n=51  0.50  (0.04 – 1.85) | n=41  0.49  (0 – 1.88) |

**New Zealand**

| **Inflammatory phenotype** | **eosinophilic** | **mixed granulocytic** | **neutrophilic** | **paucigranulocytic** | Asthmatics without sputum result | **Controls** (with sputum results) |
| --- | --- | --- | --- | --- | --- | --- |
|  | **99** | **5** | **14** | **89** | **28** | **104** |
| Female (%) | 45 (45%) | 2 (40%) | 10 (71%) | 46 (52%) | 10 (36%) | 62 (60%) |
| Age at questionnaire, years: mean (range) | 14.0 (9.1 – 20.0) | 11.8 (8.6 – 16.0) | 11.8 (8.8 – 15.7) | 15.1 (9.3 – 20.3) | 12.6 (9.0 – 17.9) | 14.9 (9.0 – 18.8) |
| Asthma diagnosis confirmed by doctor | 94 (95%) | 5 (100%) | 13 (93%) | 77 (87%) | 25 (89%) | - |
| Age at asthma diagnosis, years: median (range) | 3 (0 – 13) | 2 (1 – 4) | 4 (1 – 8) | 5 (1 – 16) | 3 (1 – 11) | - |
| missing | 20 | 0 | 2 | 19 | 4 |  |
| **Asthma severity in past 12 months*** |  |  |  |  |  | - |
| mild or moderate | 45 | 1 | 7 | 56 | 14 |  |
| severe | 54 (55%) | 4 (80%) | 7 (50%) | 33 (37%) | 14 (50%) |  |
| missing |  |  |  |  |  |  |
| **Severe asthma** (>12 attacks in past 12 months) | 18 (18%) | 1 (20%) | 2 (14%) | 8 (9%) | 7 (25%) |  |
| **Asthma medication in past 12 months** |  |  |  |  |  | - |
| none | 2 (2%) | 0 | 0 | **11 (12%)** | 1 (4%) |  |
| ICS (preventer inhaler) | **72 (73%)** | 5 (100%) | 9 (64%) | 56 (63%) | **20 (71%)** |  |
| Bronchodilator (reliever inhaler) | **91 (92%)** | 5 (100%) | 14 (100%) | 74 (83%) | **26 (93%)** |  |
| **ACQ score (past week)** |  |  |  |  |  | - |
| Median (IQR, range) |  |  |  |  |  |  |
| Well controlled (score<1.5) | 67 (76%) | 2 (40%) | 14 (100%) | 66 (84%) | 21 (88%) |  |
| Not well controlled (score ≥1.5) | 21 (24%) | 3 (60%) | 0 | 13 (16%) | 3 (12%) |  |
| missing | 11 | 0 | 0 | 10 | 4 |  |
| **Lung function** absolute values (L) & GLI-2012 z-scores | n=99 | n=5 | n=14 | n=89 | n=27 | n=104 |
| - FEV_1_, mean (SD) range | 2.65 (0.88)  1.13 – 4.84 | 1.78 (0.44)  1.20 – 2.11 | 2.14 (0.61)  1.46 – 3.34 | 3.18 (0.94)  1.36 – 5.37 | 2.29 (0.75)  1.41 – 4.27 | 3.12 (0.97)  1.31 – 5.42 |
| - FEV_1_ z-score, mean (SD) range | -0.72 (0.98)  -2.95 – 1.61 | -0.87 (1.77)  -3.52 – 0.89 | -0.28 (1.07)  -1.94 – 1.51 | -0.21 (0.97)  -2.24 – 2.36 | -0.58 (1.37)  -3.08 – 2.13 | -0.12 (0.99)  -2.41 – 2.60 |
| - FVC, mean (SD) range | 3.30 (1.10)  1.62 – 6.05 | 2.45 (0.81)  1.36 – 3.62 | 2.56 (0.69)  1.80 – 3.64 | 3.81 (1.09)  1.81 – 6.05 | 2.94 (1.09)  1.79 – 5.78 | 3.63 (1.14)  1.44 – 6.31 |
| - FVC z-score, mean (SD) range | -0.07 (0.95)  -2.15 – 2.15 | 0.35 (1.17)  -1.42 – 1.24 | 0.23 (1.35)  -2.07 – 2.58 | 0.19 (0.86)  -2.56 – 2.57 | 0.19 (1.21)  -1.62 – 2.97 | -0.01 (0.89)  -2.16 – 2.28 |
| - FEV_1_/FVC, mean (SD) range | 0.82 (0.07)  0.59 – 0.99 | 0.76 (0.13)  0.60 – 0.88 | 0.83 (0.05)  0.77 – 0.92 | 0.84 (0.07)  0.68 – 0.99 | 0.79 (0.07)  0.62 – 0.90 | 0.87 (0.06)  0.67 – 0.99 |
| - FEV_1_/FVC z-score, mean (SD) range | -0.94 (0.99)  -3.62 – 2.29 | -1.81 (1.49)  -3.59 – -0.51 | -0.87 (0.75)  -1.50 – 0.51 | -0.49 (1.07)  -2.70 – 2.28 | -1.25 (0.97)  -3.00 – 0.69 | -0.11 (0.89)  -2.68 – 2.14 |
| **FeNO level** |  |  |  |  |  |  |
| normal | 31 | 2 | 8 | 69 | 13 | 88 |
| elevated | 68 (69%) | 3 (60%) | 6 (43%) | 19 (22%) | 15 (54%) | 15 (15%) |
| not measured | 0 | 0 | 0 | 1 | 0 | 1 |
| **Skin prick test positive** | 83 (85%) | 4 (80%) | 11 (79%) | 67 (75%) | 22 (79%) | 40 (39%) |
| Not done | 1 | 0 | 0 | 0 | 0 | 1 |
| **Blood eosinophils** absolute values (10^9^/L) median (range) | n=73  0.60 (0 – 1.90) | n=4  0.60 (0.10 – 1.00) | n=9  0.50 (0.10 – 0.70) | n=77  0.30 (0 – 1.90) | n=20  0.45 (0.10 – 1.10) | n=88  0.20 (0.10 – 2.20) |

**Uganda**

| **Inflammatory phenotype** | **eosinophilic** | **mixed granulocytic** | **neutrophilic** | **paucigranulocytic** | **Asthmatics without sputum result** | **Controls** (with sputum results) |
| --- | --- | --- | --- | --- | --- | --- |
|  | **25** | **8** | **34** | **31** | **109** | **20** |
| Female (%) | 14 (56%) | 6 (75%) | 26 (76%) | 27 (87%) | 82 (75%) | 12 (60%) |
| Age at questionnaire, years: mean (range) | 15.2 (12.7 – 17.9) | 15.7 (13.0 – 17.5) | 15.5 (12.0 – 17.8) | 15.6 (12.1 – 17.9) | 15.7 (12.2 – 18.0) | 15.7 (13.0 – 18.9) |
| Asthma diagnosis confirmed by doctor | 17 (68%) | 6 (75%) | 16 (47%) | 19 (61%) | 83 (76%) | - |
| Age at asthma diagnosis, years: median (range) | 9, 1-13 | 5, 1-13 | 8, 1-16 | 12, 1-15 | 8, 1-17 | - |
| **Asthma severity in past 12 months*** |  |  |  |  |  | - |
| mild or moderate | 9 | 2 | 18 | 14 | 28 |  |
| severe | 16 (64%) | 6 (75%) | 16 (47%) | 17 (55%) | 81 (74%) |  |
| **Severe asthma** (>12 attacks in past 12 months) | 5 (20%) | 1 (13%) | 6 (18%) | 5 (16%) | 22 (20%) |  |
| **Asthma medication in past 12 months** |  |  |  |  |  | - |
| none | 6 (24%) | 2 (25%) | 12 (35%) | 10 (32%) | 25 (23%) |  |
| ICS (preventer inhaler) | 4 (16%) | 2 (25%) | 2 (6%) | 3 (10%) | 22 (20%) |  |
| Bronchodilator (reliever inhaler) | 8 (32%) | 5 (63%) | 6 (18%) | 12 (39%) | 51 (47%) |  |
| **ACQ score (past week)** |  |  |  |  |  |  |
| Median (IQR, range) | 0.5, 0 – 1.67, 0 – 2.67 | 1.0, 0 – 1.92, 0 – 2.5 | 1.1, 0.17 – 1.5, 0 – 4 | 0.33, 0 – 1.5, 0 - 3 | 0.42, 0 – 1.6, 0 – 5 | - |
| Well controlled (score<1.5) | 17 (74%) | 5 (63%) | 22 (65%) | 22 (71%) | 53 (74%) |  |
| Not well controlled (score ≥1.5) | 6 (26%) | 3 (37%) | 12 (35%) | 9 (29%) | 19 (26%) |  |
| Not done | 2 | 0 | 0 | 0 | 37 |  |
| **Lung function** absolute values (L) & GLI-2012 z-scores | n=17 | n=7 | n=27 | n=24 | n=63 | n=16 |
| - FEV_1_, mean (SD) range | 2.54 (0.60)  1.81 – 3.98 | 2.57 (1.00)  1.73 – 4.02 | 2.53 (0.35)  1.95 – 3.14 | 2.54 (0.38)  2.04 – 3.38 | 2.67 (0.45)  1.68 – 4.06 | 2.47 (0.63)  1.57 – 3.72 |
| - FEV_1_ z-score, mean (SD) range | -0.40 (0.81)  -1.48 – 1.66 | -0.18 (1.35)  -1.58 – 1.89 | -0.29 (0.76)  -1.50 – 1.03 | -0.06 (0.70)  -1.23 – 1.30 | -0.19 (0.98)  -2.78 – 2.23 | -0.26 (1.04)  -1.97 – 1.60 |
| - FVC, mean (SD) range | 2.99 (0.55)  2.34 – 4.13 | 3.55 (2.19)  2.07 – 8.01 | 2.91 (0.35)  2.25 – 3.55 | 2.92 (0.53)  2.15 – 4.57 | 3.12 (0.58)  1.77 – 5.01 | 2.89 (0.78)  1.65 – 4.53 |
| - FVC z-score, mean (SD) range | -0.03 (0.88)  -1.31 – 1.64 | 1.22 (2.91)  -1.11 – 7.14 | -0.11 (0.78)  -1.72 – 1.13 | 0.17 (1.07)  -1.82 – 2.98 | 0.07 (1.04)  -1.94 – 3.05 | 0.11 (1.52)  -2.34 – 4.13 |
| - FEV_1_/FVC, mean (SD) range | 0.85 (0.09)  0.69 – 0.99 | 0.79 (0.14)  0.49 – 0.91 | 0.87 (0.06)  0.74 – 0.97 | 0.88 (0.08)  0.67 – 0.99 | 0.86 (0.09)  0.63 – 1.00 | 0.87 (0.12)  0.48 – 1.00 |
| - FEV_1_/FVC z-score, mean (SD) range | -0.55 (1.41)  -2.69 – 1.98 | -1.45 (1.38)  -4.25 - -0.04 | -0.34 (1.00)  -2.13 – 1.46 | -0.25 (1.21)  -2.90 – 1.78 | -0.35 (1.37)  -3.14 – 2.51 | -0.26 (1.46)  -3.92 – 1.93 |
| **FeNO level** |  |  |  |  |  |  |
| normal | 6 | 2 | 22 | 24 | 59 | 18 |
| elevated | 17 (74%) | 6 (75%) | 12 (35%) | 6 (20%) | 43 (42%) | 2 (10%) |
| not measured | 2 | 0 | 0 | 1 | 7 | 0 |
| **Skin prick test positive** | 21 (84%) | 4 (50%) | 18 (53%) | 9 (30%) | 51 (52%) | 3 (15%) |
| Not tested | 0 | 0 | 0 | 1 | 10 | 0 |
| **Blood eosinophils** absolute values (10^9^/L) median (range) | n=25  0.47  (0.07 – 2.99) | n=8  0.31  (0.19 – 0.80) | n=34  0.22  (0.01 – 2.95) | n=31  0.23  (0 – 1.09) | n=98  0.22  (0.02 – 1.55) | n=20  0.26  (0.05 – 0.81) |

**United Kingdom**

| **Inflammatory phenotype** | **eosinophilic** | **mixed granulocytic** | **neutrophilic** | **paucigranulocytic** | **Asthmatics without sputum result** | **Controls** (with sputum results) |
| --- | --- | --- | --- | --- | --- | --- |
|  | **23** | **2** | **6** | **45** | **100** | **29** |
| Female (%) | 14 (61%) | 2 (100%) | 5 (83%) | 36 (80%) | 74 (74%) | 17 (59%) |
| Age at questionnaire, years: mean (range) | 25.9 (25.0 – 26.6) | 25.8 (25.6 – 26.1) | 26.0 (25.3 – 26.7) | 25.9 (24.9 – 26.9) | 25.9 (24.6 – 27.3) | 25.9 (24.7 – 27.3) |
| Asthma diagnosis confirmed by doctor | 22 (96%) | 2 (100%) | 6 (100%) | 44 (98%) | 97 (97%) | - |
| **Asthma severity in past 12 months*** |  |  |  |  |  | - |
| mild or moderate | 7 | 1 | 5 | 29 | 55 |  |
| severe | 16 (70%) | 1 (50%) | 1 (17%) | 16 (36%) | 45 (45%) |  |
| **Severe asthma** (>12 attacks in past 12 months) | 8 (35%) | 1 (50%) | 0 | 3 (7%) | 15 (15%) |  |
| **Asthma medication in past 12 months** |  |  |  |  |  | - |
| none | 1 (6%) | 0 | 0 | 4 (9%) | 22 (22%) |  |
| ICS (preventer inhaler) | 17 (74%) | 2 (100%) | 5 (83%) | 26 (58%) | 44 (44%) |  |
| Bronchodilator (reliever inhaler) | 22 (96%) | 2 (100%) | 6 (100%) | 40 (89%) | 75 (75%) |  |
| **ACQ score (past week)** |  |  |  |  |  | - |
| Median (IQR, range) | 0.92 (0.50 – 1.17, 0 – 3) | 1.0 (0.83 – 1.17, 0.83 – 1.17) | 0.25 (0 – 0.67, 0 – 1.33) | 0.33 (0 – 1.17, 0 – 2.5) | 0.17 (0 – 0.83, 0 – 2.83) |  |
| Well controlled (score<1.5) | 19 (86%) | 2 (100%) | 6 (100%) | 38 (86%) | 89 (92%) |  |
| Not well controlled (score ≥1.5) | 3 (14%) | 0 | 0 | 6 (14%) | 8 (8%) |  |
| Not done | 1 | 0 | 0 | 1 | 3 |  |
| **Lung function** absolute values (L) & GLI-2012 z-scores | N=23 | N=2 | N=5 | N=44 | N=96 | N=29 |
| - FEV_1_, mean (SD) range | 3.70 (0.90)  2.39 – 5.45 | 2.95 (0.95)  2.28 – 3.62 | 3.77 (0.75)  2.97 – 4.62 | 3.33 (0.78)  1.62 – 5.29 | 3.50 (0.74)  2.36 – 5.61 | 3.98 (0.90)  2.59 – 6.32 |
| - FEV_1_ z-score, mean (SD) range | -0.33 (0.91)  -2.26 – 0.96 | -0.88 (3.02)  -3.02 – 1.26 | -0.07 (1.60)  -1.64 – 2.43 | -0.56 (1.43)  -4.12 – 3.59 | -0.36 (1.00)  -3.02 – 2.82 | -0.15 (1.05)  -2.62 – 2.76 |
| - FVC, mean (SD) range | 4.75 (1.23)  3.33 – 7.78 | 4.07 (1.24)  3.19 – 4.95 | 4.53 (0.82)  3.65 – 5.69 | 4.05 (0.95)  1.80 – 6.34 | 4.27 (1.03)  2.78 – 7.45 | 4.81 (1.31)  3.31 – 8.11 |
| - FVC z-score, mean (SD) range | 0.31 (0.85)  -1.62 – 1.96 | 0.35 (3.33)  -2.00 – 2.70 | 0.02 (1.26)  -1.61 – 1.67 | -0.33 (1.32)  -4.52 – 3.74 | -0.13 (0.89)  -2.03 – 1.65 | -0.06 (1.10)  -2.12 – 3.12 |
| - FEV_1_/FVC, mean (SD) range | 0.79 (0.08)  0.58 – 0.92 | 0.72 (0.01)  0.71 – 0.73 | 0.83 (0.04)  0.80 – 0.90 | 0.83 (0.07)  0.67 – 0.95 | 0.83 (0.08)  0.47 – 0.97 | 0.84 (0.06)  0.70 – 0.97 |
| - FEV_1_/FVC z-score, mean (SD) range | -0.88 (1.09)  -3.18 – 1.00 | -1.90 (0.11)  -1.97 – -1.82 | -0.33 (0.64)  -0.89 – 0.73 | -0.42 (1.00)  -2.73 – 1.71 | -0.34 (1.06)  -4.05 – 2.20 | -0.15 (0.91)  -2.11 – 1.96 |
| **Skin prick test positive** | 17 (94%) | 2 (100%) | 3 (50%) | 27 (73%) | 75 (84%) | 9 (33%) |
| Not tested | 5 | 0 | 0 | 8 | 11 | 2 |
| **Blood eosinophils** absolute values (10^9^/L) median (range) | n=19  0.40 (0.11 – 0.81) | n=2  0.22 (0.06 – 0.37) | n=5  0.14 (0.09 – 0.56) | n=34  0.15 (0.02 – 0.41) | n=65  0.19 (0.02 – 0.83) | n=26  0.11 (0.02 – 0.51) |

**Online appendix 2: Comparison of sputum slide results, excluding low quality slides***

| **Centre** | **Brazil** | **Brazil excluding low quality slides*** | **Ecuador** | **Ecuador excluding low quality slides*** | **New Zealand** | **New Zealand, excluding low quality slides*** | **Uganda** | **Uganda excluding low quality slides*** | **United Kingdom** | **United Kingdom**  **excluding low quality slides*** |
| --- | --- | --- | --- | --- | --- | --- | --- | --- | --- | --- |
| Sputum inflammatory phenotype: **asthma** **cases** | N=115 | N=87 | N=125 | N=111 | N=207 | N=116 | N=98 | N=75 | N=76 | N=49 |
| eosinophilic | 38 (33%) | 26 (30%) | 35 (28%) | 31 (28%) | 99 (48%) | 57 (49%) | 25 (25%) | 15 (20%) | 23 (30%) | 14 (29%) |
| mixed granulocytic | 2 (2%) | 2 (2%) | 5 (4%) | 5 (5%) | 5 (2%) | 3 (3%) | 8 (8%) | 7 (9%) | 2 (3%) | 2 (4%) |
| neutrophilic | 5 (4%) | 5 (6%) | 8 (6%) | 7 (6%) | 14 (7%) | 11 (9%) | 34 (35%) | 30 (40%) | 6 (8%) | 6 (12%) |
| paucigranulocytic | 70 (61%) | 54 (62%) | 77 (62%) | 68 (61%) | 89 (43%) | 45 (39%) | 31 (32%) | 23 (31%) | 45 (59%) | 27 (55%) |
| **Repeat sputum slide** |  |  |  |  |  |  |  |  |  |  |
| same phenotype (EA or NEA**) | 27 (68%) | 22 (69%) | 25 (69%) | 24 (69%) | 72 (67%) | 41 (64%) | 9 (75%) | 9 (90%) | 5 (56%) | 3 (60%) |
| Changed: |  |  |  |  |  |  |  |  |  |  |
| EA to NEA | 6 | 6 | 4 | 4 | 18 | 12 | 0 | 0 | 3 | 1 |
| NEA to EA | 7 | 4 | 7 | 7 | 17 | 11 | 3 | 1 | 1 | 1 |
| **Controls** | N=20 | N=11 | N=41 | N=39 | N=104 | N=64 | N=20 | N=17 | N=29 | N=22 |
| eosinophilic | 4 (20%) | 2 (18%) | 3 (7%) | 3 (8%) | 11 (11%) | 8 (12%) | 2 (10%) | 1 (6%) | 3 (10%) | 2 (9%) |
| mixed granulocytic | 0 | 0 | 0 | 0 | 1 (1%) | 1 (2%) | 1 (5%) | 1 (6%) | 0 | 0 |
| neutrophilic | 4 (20%) | 3 (27%) | 1 (2%) | 1 (2%) | 11 (11%) | 6 (9%) | 12 (60%) | 10 (59%) | 3 (10%) | 3 (14%) |
| paucigranulocytic | 12 (60%) | 6 (55%) | 37 (90%) | 35 (90%) | 81 (78%) | 49 (77%) | 5 (25%) | 5 (29%) | 23 (79%) | 17 (77%) |

*<400 total squamous cells and ≥30% squamous cells. ** EA (eosinophilic or mixed); NEA (neutrophilic or paucigranulocytic)

**Online appendix 3** Sputum slide results with original and alternative phenotype definitions: A) 1% cut-off for eosinophils^1^ and B) 54% cut-off for neutrophils^2^

| **Centre** | **Brazil** | **Brazil A** | **Brazil B** | **Ecuador** | **Ecuador A** | **Ecuador B** | **NZ** | **NZ**  **A** | **NZ**  **B** | **Uganda** | **Uganda**  **A** | **Uganda**  **B** | **UK** | **UK**  **A** | **UK**  **B** |
| --- | --- | --- | --- | --- | --- | --- | --- | --- | --- | --- | --- | --- | --- | --- | --- |
| Sputum inflammatory phenotype: **asthma** **cases** | N=115 | N=115 | N=115 | N=125 | N=125 | N=125 | N=207 | N=207 | N=207 | N=98 | N=98 | N=98 | N=76 | N=76 | N=76 |
| eosinophilic | 38 (33%) | 56 (49%) | 38 (33%) | 35 (28%) | 43 (34%) | 35 (28%) | 99 (48%) | 130 (63%) | 94 (45%) | 25 (25%) | 30 (31%) | 24 (24%) | 23 (30%) | 39 (51%) | 22 (29%) |
| mixed granulocytic | 2 (2%) | 2 (2%) | 2 (2%) | 5 (4%) | 7 (6%) | 5 (4%) | 5 (2%) | 12 (6%) | 10 (5%) | 8 (8%) | 16 (16%) | 9 (9%) | 2 (3%) | 4 (5%) | 3 (4%) |
| neutrophilic | 5 (4%) | 5 (4%) | 8 (7%) | 8 (6%) | 6 (5%) | 9 (7%) | 14 (7%) | 7 (3%) | 17 (8%) | 34 (35%) | 26 (27%) | 39 (40%) | 6 (8%) | 4 (5%) | 11 (14%) |
| paucigranulocytic | 70 (61%) | 52 (45%) | 67 (58%) | 77 (62%) | 69 (55%) | 76 (61%) | 89 (43%) | 58 (28%) | 86 (42%) | 31 (32%) | 26 (27%) | 26 (27%) | 45 (59%) | 29 (38%) | 40 (53%) |
| **Repeat sputum slide** |  |  |  |  |  |  |  |  |  |  |  |  |  |  |  |
| same phenotype (EA or NEA**) | 27 (68%) | 29 (73%) | 27 (68%) | 25 (69%) | 29 (81%) | 25 (69%) | 72 (67%) | 72 (67%) | 72 (67%) | 9 (75%) | 10 (83%) | 9 (75%) | 5 (56%) | 5 (56%) | 5 (56%) |
| Changed: |  |  |  |  |  |  |  |  |  |  |  |  |  |  |  |
| EA to NEA | 6 | 4 | 6 | 4 | 2 | 4 | 18 | 19 | 18 | 0 | 0 | 0 | 3 | 3 | 3 |
| NEA to EA | 7 | 7 | 7 | 7 | 5 | 7 | 17 | 16 | 17 | 3 | 2 | 3 | 1 | 1 | 1 |
| **Controls** | N=20 | N=20 | N=20 | N=41 | N=41 | N=41 | N=104 | N=104 | N=104 | N=20 | N=20 | N=20 | N=29 | N=29 | N=29 |
| eosinophilic | 4 (20%) | 5 (25%) | 4 (20%) | 3 (7%) | 8 (20%) | 3 (7%) | 11 (11%) | 24 (23%) | 10 (10%) | 2 (10%) | 2 (10%) | 2 (10%) | 3 (10%) | 6 (21%) | 3 (10%) |
| mixed granulocytic | 0 | 0 | 0 | 0 | 0 | 0 | 1 (1%) | 1 (1%) | 2 (2%) | 1 (5%) | 6 (30%) | 1 (5%) | 0 | 0 | 0 |
| neutrophilic | 4 (20%) | 4 (20%) | 5 (25%) | 1 (2%) | 1 (2%) | 1 (2%) | 11 (11%) | 11 (11%) | 17 (16%) | 12 (60%) | 7 (35%) | 13 (65%) | 3 (10%) | 3 (10%) | 3 (10%) |
| paucigranulocytic | 12 (60%) | 11 (55%) | 11 (55)% | 37 (90%) | 32 (78%) | 37 (90%) | 81 (78%) | 68 (65%) | 75 (72%) | 5 (25%) | 5 (25%) | 4 (20%) | 23 (79%) | 20 (69%) | 23 (79%) |

^1^ eosinophilic: ≥1% eosinophils; neutrophilic: <1% eosinophils and ≥61% neutrophils; mixed granulocytic: ≥1% eosinophils and ≥61% neutrophils; paucigranulocytic: <1% eosinophils and <61% neutrophils.

^2^ eosinophilic: ≥2.5% eosinophils; neutrophilic: <2.5% eosinophils and ≥54% neutrophils; mixed granulocytic: ≥2.5% eosinophils and ≥54% neutrophils; paucigranulocytic: <2.5% eosinophils and <54% neutrophils.

**Online supplementary information**

Ethical approval for the study has been obtained from the LSHTM ethics committee (ref: 9776) and in all five study centres. Informed consent was obtained from all participants or their parents/carers before taking part.

ALSPAC: Ethical approval for the UK-arm of the study was obtained from the ALSPAC Ethics and Law Committee, and the Local Research Ethics Committees. Consent for biological samples has been collected in accordance with the Human Tissue Act (2004). REDcap was used to collect the ALSPAC data (<http://projectredcap.org/resources/citations/>). Informed consent for the use of data collected via questionnaires and clinics was obtained from participants following the recommendations of the ALSPAC Ethics and Law Committee at the time. The ALSPAC study website contains details of all the data that is available through a fully searchable data dictionary and variable search tool (http://www.bristol.ac.uk/alspac/researchers/our-data/)/.
